# Evaluating the Reproducibility and Verifiability of Nutrition Research: A Case Study of Studies Assessing the Relationship Between Potatoes and Colorectal Cancer

**DOI:** 10.1101/2024.12.01.24318272

**Authors:** Yasaman Jamshidi-Naeini, Colby J. Vorland, Pranav Kapoor, Bailey Ortyl, Jocelyn Mineo, Luke Still, Korlu Sorsor, Shelby Rodney, Xander Tooze, Brent Flickinger, Beate Henschel, Stephanie L. Dickinson, David B. Allison

## Abstract

**Background:** The credibility of nutritional research is dependent on the rigor with which studies are conducted and the ability for independent assessment to be performed. Despite the importance of these, more work is needed in the field of nutrition to buttress the trustworthiness of nutrition research.

**Objective:** To develop and apply a process for evaluating the rigor, reproducibility, and verifiability of nutritional research, using the relationship between potato consumption and Colorectal cancer (CRC) as a case study.

**Methods:** We updated existing systematic reviews to include studies on potatoes and CRC, assessing their design, execution, and reporting quality. We attempted to reproduce and verify the results of included studies by requesting raw data from authors and following statistical methods as described in the publications. Rigor was evaluated using four different tools: ROBINS-E, STROBE-Nut, Newcastle-Ottawa scale, and additional criteria related to transparency.

**Results:** Eighteen studies were included, none of which publicly share data. We managed to access data for only two studies, successfully reproducing and verifying the results for one. The majority of studies exhibited a high risk of bias, with significant limitations in reporting quality and methodological rigor.

**Conclusions:** Research on the relationship between potato consumption and CRC risk is insufficiently reproducible and verifiable, undermining the trustworthiness of its findings. This study highlights the need for improving transparency, data sharing, and methodological rigor in nutritional research. Our approach provides a model for assessing the credibility of research in other areas of nutrition.

## Introduction

Trust in nutrition science appears to be lower than is trust in other domains of science (1, 2).Trust in science should be predicated on the *trustworthiness* of the science (3). In turn, the trustworthiness of nutritional research relies in part on the independent assessment of the quality of the research. This depends on how well the research is designed, executed, analyzed, and reported (i.e., rigor^1^). Trustworthiness can be enhanced if raw data can be obtained and independently analyzed to assess reproducibility^2^ and verifiability^3^. Other research has examined reproducibility of research published in specific journals (6–12) or disciplines (13, 14). To our knowledge, no study has evaluated research on a nutrition topic for reproducibility, nor evaluated the verifiability.

Potatoes are a staple food for many populations worldwide, estimated to be the third most consumed crop (15). As such, it is paramount to understand their relationship with health. The Scientific Report of the 2020 Dietary Guidelines Advisory Committee assigned a grade of ‘moderate evidence’ that dietary patterns higher in foods including potatoes are associated with a greater colon and rectal cancer (CRC) risk (16). However, it is not clear from the report whether potatoes in isolation are related to CRC risk. While some systematic reviews have been published on the topic (17, 18), traditional systematic review procedures do not evaluate reproducibility and verifiability of research on a particular research question. Herein, we develop a process to assess the rigor, reproducibility, and verifiability of nutritional research, using the relationship between potatoes and CRC as a case study.

## Methods

### Search Strategy and Screening

We identified three recent systematic reviews by Schwingshackl et al. (2018) (17), Mofrad et al. (2021) (18), and Mofrad et al. 2020 (19) that drew conclusions about the association between potatoes and CRC risk. We also updated the systematic review of (17) up to June 2022 to identify new studies since their last search. We first modified their PubMed search string based on the purpose of the current work, and then with the help of a librarian, constructed our search strings for Web of Science and Scopus (**Appendix 1**). The search was run on June 13, 2022. Two analysts independently screened all studies for relevance and included all that met predefined inclusion and exclusion criteria using Covidence (**Figure 1**). We included studies that met the following criteria: Study design (self-identified by the authors): cohort, case–control, nested case–control, or randomized controlled trial; human participants aged 18 years or older on average; participants free of CRC at the onset of the study unless specified otherwise; studies on CRC recurrence or in CRC survivors were included if participants were not currently diagnosed with cancer, and for case-control designs, studies were included if there was a control group without CRC, even if the “cases” had CRC; studies must have included an analysis for the independent relationship between potatoes (in any form) and CRC. We excluded studies that met the following criteria: study type: reviews, animal studies, and in vitro studies; participants: studies including participants younger than 18 years of age on average or exclusively pregnant women; studies where all participants had CRC at the onset, except in case-control studies where this criterion did not apply; studies without an independent analysis for the relationship between potatoes (in any form) and CRC. Characteristics were extracted from each study, including the design, cohort used, follow-up length, statistical methods and adjustment factors, sample size, country, gender(s) studied, dietary and cancer assessment methods, stage of cancer for the participants, potato type studied, inclusion and exclusion criteria, results, funding, data sharing statements, and the current journal data sharing policy.

**Figure 1.**
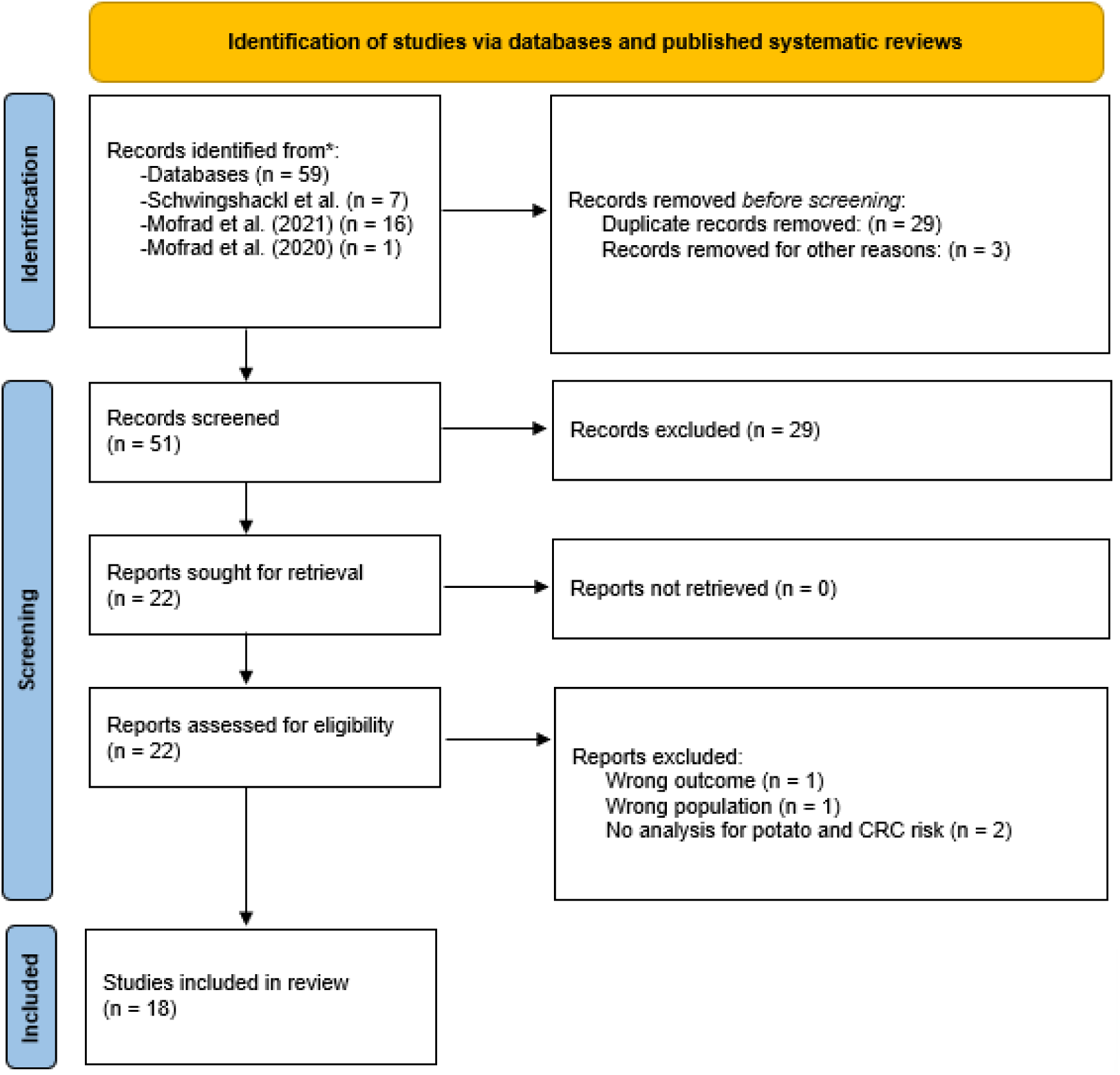
**Flowchart of study identification and screening process.**

### Assessing Reproducibility and Verifiability

We first attempted to contact authors and request raw data by emailing, mailing letters, and calling them. We first emailed the corresponding author, and, if upon not receiving a reply, emailed other authors whereby we could locate them based on name and affiliation listed in the publication.

Where we could obtain access to raw data, we first assessed whether we could obtain the same results for the potato-CRC association(s) by following the methods as published (**Figure 2**). We first checked whether, following the reported methods in the article, we could obtain the same number of participants included in the potato-CRC association(s) and whether the reported summary statistics could be reproduced. Each check was conducted by a statistician member of our team who documented their process in statistical code, and the process was checked by a senior statistician. For any articles that could not be reproduced, we emailed the author(s) and shared our misunderstandings on our part.

**Figure 2.**
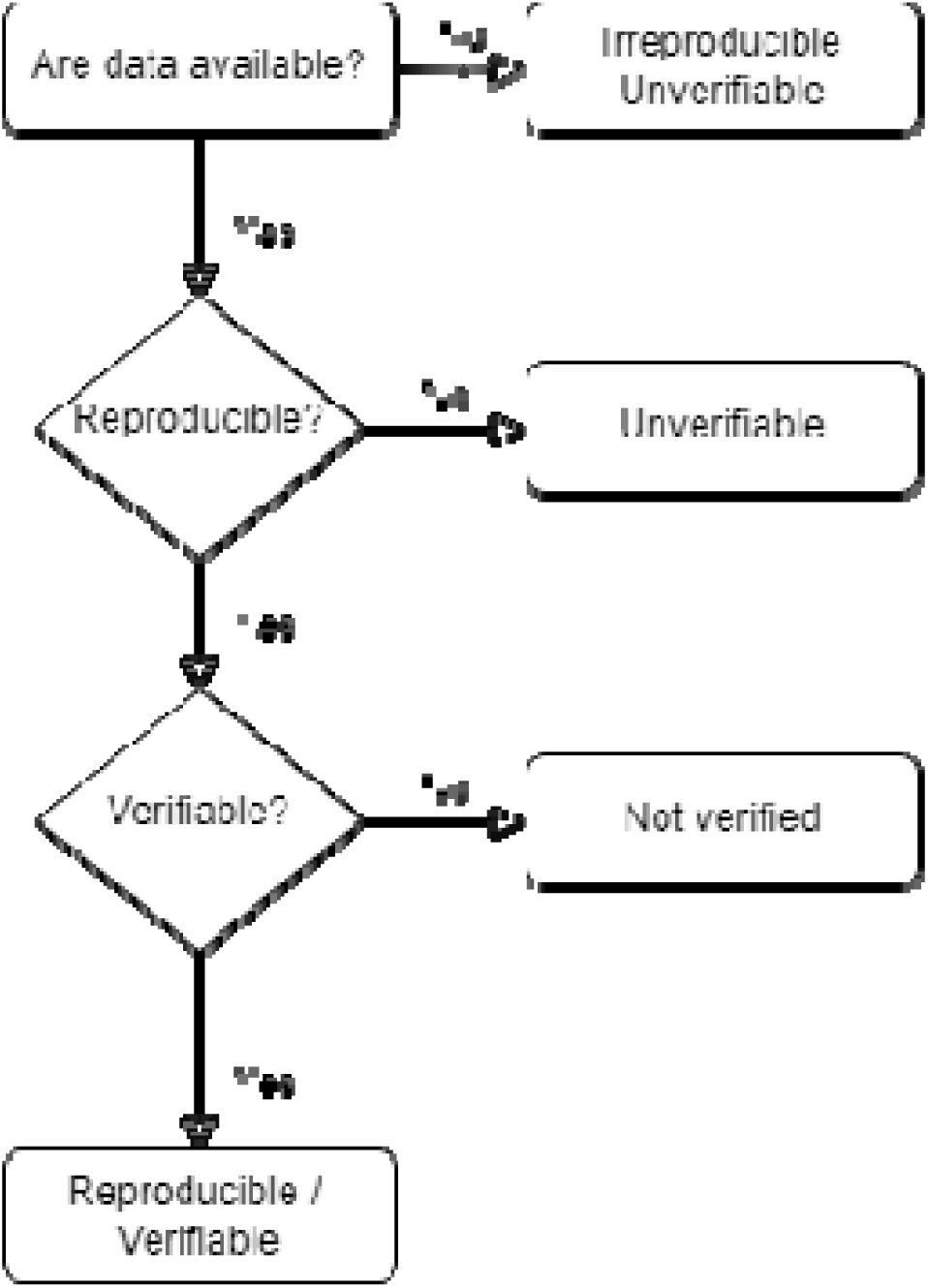
**Flowchart of reproducibility and verifiability assessment.**

If, after those procedures, we could not obtain the same results, or if the authors did not respond, the results were considered irreproducible, and thus unverifiable. If we could reproduce the results, we verified the results by assessing whether the statistical methods were appropriate for the study design and whether the statistical conclusions

### Assessment of rigor

To assess the rigor of the studies included, we used four methods. For each method, two investigators independently rated each article, and discrepancies were resolved through discussion.

- **ROBINS-E tool**, which assesses risk of bias in non-randomized studies of exposures (20).
- **STROBE-Nut checklist**, for reporting quality with an extension for nutritional epidemiology (21).
- **Newcastle-Ottawa scale**, a quality checklist for non-randomized studies (22).
- **Additional criteria** that we specified related to indicators of rigor (registration of in scientific articles identified in our previous work (23).

The IRB of Indiana University determined that this study did not require human subjects review.

## Results

Eighteen studies were included (24–41); their characteristics are described in **Table 1**. All studies were non-randomized, which limits the ability to draw causal conclusions. Of the 18 studies, 11 were prospective cohorts and 7 were case controls. The studies were done in 12 different countries. Six studied included only women, the others included both women and men. Seventeen studies used a food-frequency questionnaire to assess potato consumption, and one used 24-hour dietary recalls. CRC was rigorously ascertained in almost all studies using registry linkage or review of medical records. None of the studies publicly shared data, and two studies indicated that data were available on request.

**Table 1.** Characteristics of Included Studies.

| Study | Design | Country | Gender(s) Studied | Dietary Assessment Method | Potato Description as the Exposure | Outcome Ascertainment |
| --- | --- | --- | --- | --- | --- | --- |
| **Asli 2016** | Prospective cohort | Norway | Women | FFQ | Potatoes (details unspecified) | Registry linkage |
| **Flood 2002** | Prospective cohort | USA | Women | FFQ | Potatoes excluding French fries | Registry linkage |
| **Kato 1997** | Prospective cohort | USA | Women | FFQ | Potatoes (details unspecified) | Self-report & medical records |
| **Michels 2000** | Prospective cohort | USA | Women, Men | FFQ | Potatoes (mashed or baked), French fries, Potato chips | Self-report & medical records |
| **Steinmetz 1994** | Prospective cohort | USA | Women | FFQ | Potatoes (details unspecified) | Registry linkage |
| **Mucci 2006** | Prospective cohort | Sweden | Women | FFQ | Pan-fried potatoes, Potato chips and French fries | Registry linkage |
| **Yeh 2006** | Prospective cohort | Taiwan | Women, Men | FFQ | Sweet potatoes | Registry linkage & medical records |
| **Liu 2021** | Case-control | China | Women, Men | FFQ | Potatoes (details unspecified) | Medical records |
| **Partula 2020** | Prospective cohort | France | Women, Men | 24-h recall | Potato and tuber fiber | Self-report & medical records |
| **Tayyem 2018** | Case-control | Jordan | Women, Men | FFQ | Fried potato, potato and corn chips | Medical records |
| **Agnoli 2013** | Prospective cohort | Italy | Women, Men | FFQ | Potatoes (details unspecified) | Medical records |
| **Annema 2011** | Case-control | Australia | Women, Men | FFQ | Potatoes (details unspecified) | Registry linkage |
| **Deneo-Pellegrini 1996** | Case-control | Uruguay | Women, Men | FFQ | Potatoes, sweet potatoes | Medical records |
| **Khan 2004** | Prospective cohort | Japan | Women, Men | FFQ | Potatoes (details unspecified) | Self-report or family contact |
| **Levi 1999** | Case-control | Switzerland | Women, Men | FFQ | Potatoes (details unspecified) | Medical records |
| **Lin 2005** | Prospective cohort | USA | Women | FFQ | Potatoes, French fries, Potato chips | Self-report & medical records |
| **Ping 1998** | Case-control | Japan | Women, Men | FFQ | Potato products (details unspecified) | Medical records |
| **Tayyem 2014** | Case-control | Jordan | Women, Men | FFQ | Sweet potatoes, mashed potatoes | Medical records |

### Reproducibility and verifiability assessment

We received no response from 10 of 18 research groups (56%) (**Table 2**). Two contacts indicated that they were retired and declined to further work with us. We received one cost estimate to access data that was above our available resources ($51,644.73). One group shared a data file that was insufficient to reproduce analyses and declined to share additional data. One group was willing to share data conditional on IRB approval and a data use agreement, but the timeline from contacting them to initiating these was too long (> 8 months). One group permitted (required as a condition of granting access to the data) us to visit their group in another country (France) and analyze their data on their computers, and one group sent us all data needed to reproduce and verify their analyses.

**Table 2.** Summary of Contacting Authors for Raw Data.

| **Result of Contact** | **Number of Studies** |
| --- | --- |
| No response | 10 |
| Responded but declined to share data (reason: retired) | 2 |
| Shared data file but was insufficient to reproduce analyses | 1 |
| Cost to access too high | 1 |
| Language barrier on application for data access | 1 |
| Willing to share conditional on local IRB approval and data use agreement, but timeline to initiate these processes was too long (> 8 months) | 1 |
| Allowed data access from institutional servers | 1 |
| Shared data | 1 |

Of the two studies for which we had access to data (33, 40), we could reproduce and verify the results of one study, and we could not reproduce and therefore not verify the results of the other study.

### Reproducibility analysis of Flood et al

Details and numerical values of our reproducibility results are provided in **Appendix 2.** In summary, we first aimed to reproduce the quintiles of fruit and vegetable intake as reported in Table 2 of Flood et al. (2002). While there were some discrepancies within the original paper regarding the number of participants included in the analyses (e.g., inconsistencies between the text and table footnote), we obtained a sample size nearly identical to that in the table footnote (45,490 in the paper vs. 45,494 in our analysis).

The paper reported hazard ratios (HRs) and 95% confidence intervals (95% CIs) for quintiles of fruit and vegetable intake in servings per day per 1,000 kilojoules. However, the dataset we received did not include a variable for servings per day. Without a variable for servings, we tested whether we could use frequency of consumption by dividing the weekly frequencies by seven to estimate daily servings. We then converted kilocalories to kilojoules to calculate servings per day per 1,000 kilojoules.

We obtained reasonably comparable values for median and cutoff points across the quintiles of fruit intake. However, for vegetable intake, we were unable to reproduce the median and cutoff values reported in the paper (Table 2 of the original paper). A similar pattern emerged for hazard ratios. The HRs we calculated based on the quintile cutoffs provided in the original paper were largely consistent with those reported by Flood et al. (2002) for fruit intake, but not for vegetable intake (which includes potatoes). For example, for quintile 5 of vegetable intake, we obtained an HR of 0.567, while the paper reported an HR of 0.92.

We also calculated HRs using the cutoffs derived from our analysis (which differed from those reported in the paper), as well as through additional exploratory analyses in an attempt to reproduce the original HRs. We were unable to reproduce the exact results. However, these differences did not alter the overall conclusions of the study. That is, despite likely using a different method to define servings, our results closely approximated the published results, leading to conclusions similar to those of the original paper. Given our inability to reproduce the broader category results for total vegetable intake and quintile cutoffs, we did not proceed with calculating HRs specifically for potato consumption and CRC.

The authors of the original study were collegial throughout the process, and this was the only study for which the complete dataset required for reproducing analyses was sent to us. However, with over twenty years having passed since the original study, the lead author had since moved institutions and no longer had access to all the data as they did at the time of publication. While the authors were willing to assist, the time elapsed was a limiting factor for providing further clarification. This experience highlights the importance of robust documentation of statistical procedures and data sharing to ensure the reproducibility of scientific research — an aspect of scientific research that is more widely recognized and emphasized today than it was when the original study was conducted.

### Reproducibility analysis of Partula et al

To reproduce the analyses in Partula et al., a condition of access to the data required members of our team to travel from the United States to France to work on the data on their servers. Prior to our visit, we were provided with the analytical code, which we reviewed to familiarize ourselves with the approach. Once in France, we checked and re-ran their code on the final analytic dataset for the association between potato fiber and CRC. However, we were not allowed to take the analysis output off their servers. Instead, we documented the output on-site and cross-checked the numbers against those reported in the paper. We obtained the same numerical values, with minor discrepancies in the second or third decimal places for a few values. These differences did not affect the overall conclusions of the analysis. Thus, we reproduced the models related to potato fiber intake and CRC. We also verified that the statistical procedures used were appropriate to answer the study question.

### Assessment of rigor

#### ROBINS-E

The results of the assessment of the seven domains are shown in **Table 3**. Sixteen (89%) of the studies were judged at high risk of bias, mostly due to measurement weaknesses and lack of specified or state-of-the-art missing data handling procedures.

**Table 3.**
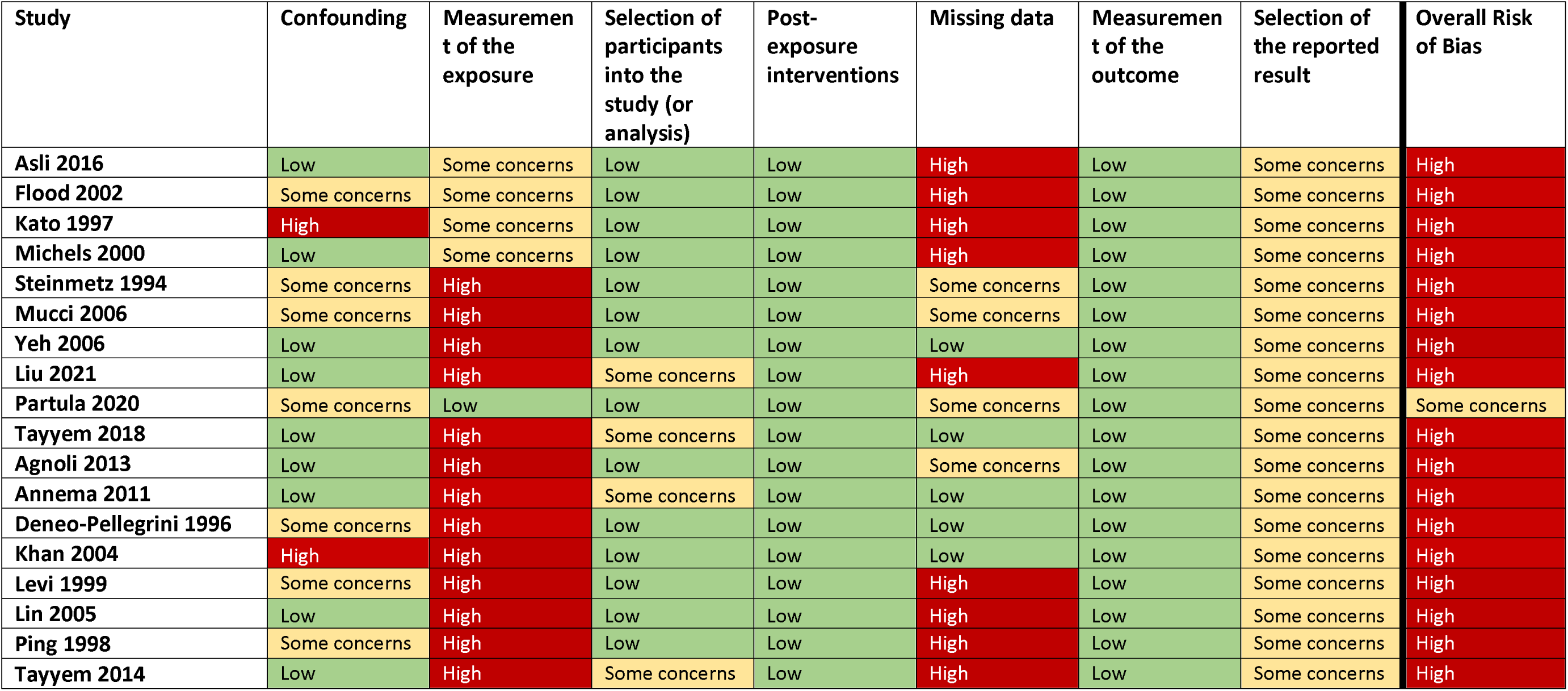
Risk of Bias Assessment Using ROBINS-E.

| Study | Confounding | Measurement of the exposure | Selection of participants into the study (or analysis) | Post-exposure interventions | Missing data | Measurement of the outcome | Selection of the reported result | Overall Risk of Bias |
| --- | --- | --- | --- | --- | --- | --- | --- | --- |
| Asli 2016 | Low | Some concerns | Low | Low | High | Low | Some concerns | High |
| Flood 2002 | Some concerns | Some concerns | Low | Low | High | Low | Some concerns | High |
| Kato 1997 | High | Some concerns | Low | Low | High | Low | Some concerns | High |
| Michels 2000 | Low | Some concerns | Low | Low | High | Low | Some concerns | High |
| Steinmetz 1994 | Some concerns | High | Low | Low | Some concerns | Low | Some concerns | High |
| Mucci 2006 | Some concerns | High | Low | Low | Some concerns | Low | Some concerns | High |
| Yeh 2006 | Low | High | Low | Low | Low | Low | Some concerns | High |
| Liu 2021 | Low | High | Some concerns | Low | High | Low | Some concerns | High |
| Partula 2020 | Some concerns | Low | Low | Low | Some concerns | Low | Some concerns | Some concerns |
| Tayyem 2018 | Low | High | Some concerns | Low | Low | Low | Some concerns | High |
| Agnoli 2013 | Low | High | Low | Low | Some concerns | Low | Some concerns | High |
| Annema 2011 | Low | High | Some concerns | Low | Low | Low | Some concerns | High |
| Deneo-Pellegrini 1996 | Some concerns | High | Low | Low | Low | Low | Some concerns | High |
| Khan 2004 | High | High | Low | Low | Low | Low | Some concerns | High |
| Levi 1999 | Some concerns | High | Low | Low | High | Low | Some concerns | High |
| Lin 2005 | Low | High | Low | Low | High | Low | Some concerns | High |
| Ping 1998 | Some concerns | High | Low | Low | High | Low | Some concerns | High |
| Tayyem 2014 | Low | High | Some concerns | Low | High | Low | Some concerns | High |

#### STROBE-Nut

The results of the assessment of the six sections are shown in **Table 4**. Most items in the ‘title and abstract’, ‘introduction’, and ‘discussion’ sections were reported, but some items tended to be missing from ‘methods’, ‘results’, and ‘other information’ sections

### Newcastle-Ottawa scale

The number of stars for each section, as well as the total stars, are shown in **Table 5**.

**Table 4.** Reporting Quality Assessed Using STROBE-Nut.

| **Study** | **Title & Abstract** | **Introduction** | **Methods** | **Results** | **Discussion** | **Other information** |
| --- | --- | --- | --- | --- | --- | --- |
| **Asli 2016** | 67% | 100% | 83% | 100% | 100% | 33% |
| **Flood 2002** | 100% | 100% | 74% | 90% | 100% | 0% |
| **Kato 1997** | 33% | 100% | 58% | 60% | 83% | 0% |
| **Michels 2000** | 67% | 100% | 65% | 91% | 100% | 0% |
| **Steinmetz 1994** | 67% | 100% | 72% | 70% | 100% | 0% |
| **Mucci 2006** | 67% | 100% | 65% | 92% | 100% | 0% |
| **Yeh 2006** | 67% | 100% | 56% | 70% | 67% | 0% |
| **Liu 2021** | 100% | 100% | 68% | 50% | 100% | 33% |
| **Partula 2020** | 100% | 100% | 85% | 92% | 100% | 67% |
| **Tayyem 2018** | 100% | 100% | 75% | 44% | 83% | 33% |
| **Agnoli 2013** | 100% | 100% | 72% | 70% | 100% | 0% |
| **Annema 2011** | 100% | 100% | 80% | 73% | 100% | 33% |
| **Deneo-Pellegrini 1996** | 100% | 100% | 70% | 44% | 100% | 0% |
| **Khan 2004** | 67% | 100% | 39% | 60% | 100% | 0% |
| **Levi 1999** | 67% | 100% | 74% | 25% | 67% | 0% |
| **Lin 2005** | 100% | 100% | 74% | 90% | 100% | 0% |
| **Ping 1998** | 67% | 50% | 70% | 44% | 50% | 0% |
| **Tayyem 2014** | 67% | 100% | 75% | 44% | 83% | 33% |

**Table 5.** Quality Assessment Using Newcastle-Ottawa Scale.

| Study | Selection (# Stars) | Comparability (# Stars) | Exposure/Outcome (# Stars) | Total Stars |
| --- | --- | --- | --- | --- |
| Asli 2016 | 3 | 2 | 3 | 8 |
| Flood 2002 | 3 | 2 | 2 | 7 |
| Kato 1997 | 3 | 2 | 3 | 8 |
| Michels 2000 | 2 | 2 | 1 | 5 |
| Steinmetz 1994 | 3 | 2 | 2 | 7 |
| Mucci 2006 | 3 | 2 | 3 | 8 |
| Yeh 2006 | 4 | 2 | 2 | 8 |
| Liu 2021 | 2 | 2 | 3 | 7 |
| Partula 2020 | 2 | 2 | 3 | 7 |
| Tayyem 2018 | 1 | 2 | 1 | 4 |
| Agnoli 2013 | 2 | 2 | 3 | 7 |
| Annema 2011 | 3 | 2 | 1 | 6 |
| Deneo-Pellegrini 1996 | 2 | 2 | 2 | 6 |
| Khan 2004 | 3 | 2 | 1 | 6 |
| Levi 1999 | 2 | 2 | 1 | 5 |
| Lin 2005 | 2 | 2 | 2 | 6 |
| Ping 1998 | 3 | 2 | 1 | 6 |
| Tayyem 2014 | 1 | 2 | 1 | 4 |

"A study is judged on three broad perspectives: the selection of the study groups; the comparability of the groups; and the ascertainment of either the exposure or outcome of interest for case-control or cohort studies respectively." Stars are awarded such that the highest quality studies are awarded up to nine stars. https://www.ohri.ca/programs/clinical_epidemiology/oxford.asp

#### Additional Criteria

The results of the assessment are reported in **Table 6**. No studies were preregistered, had publicly available data or code, or corrected for multiple testing. Six (33%) used inappropriate causal language, and three (17%) exaggerated non-significant effects. Seven (39%), based on the methods and statistical analysis described, appear to contain errors in failing to statistically account for sources of clustering that would require raw data to definitively confirm.

**Table 6.** Additional Criteria.

| Study | Pre-registered? | Data publicly available? | Code publicly available? | Inappropriate causal language? | Exaggerating significant effect<sup>2</sup> | Exaggerating non-significant effect<sup>3</sup> | Clustering errors<sup>4</sup> | DINS errors<sup>5</sup> | Controlled for multiple testing |
| --- | --- | --- | --- | --- | --- | --- | --- | --- | --- |
| Asli 2016 | No | No | No | No | No | No | No | No | No |
| Flood 2002 | No | No | No | No | No | No | No | No | No |
| Kato 1997 | No | No | No | No | No | No | No | No | No |
| Michels 2000 | No | No | No | Yes | No | No | No | No | No |
| Steinmetz 1994 | No | No | No | No | No | Yes | No | No | No |
| Mucci 2006 | No | No | No | No | No | No | Likely | No | No |
| Yeh 2006 | No | No | No | No | No | No | Likely | No | No |
| Liu 2021 | No | No | No | Yes | No | No | Likely | No | No |
| Partula 2020 | No | No | No | No | No | No | No | No | No |
| Tayyem 2018 | No | No | No | Yes | No | Yes | Likely | No | No |
| Agnoli 2013 | No | No | No | No | No | No | Likely | No | No |
| Annema 2011 | No | No | No | Yes | No | No | No | No | No |
| Deneo-Pellegrini 1996 | No | No | No | No | No | No | Likely | No | No |
| Khan 2004 | No | No | No | Yes | No | No | No | No | No |
| Levi 1999 | No | No | No | No | No | No | No | No | No |
| Lin 2005 | No | No | No | No | No | No | No | No | No |
| Ping 1998 | No | No | No | Yes | No | No | No | No | No |
| Tayyem 2014 | No | No | No | No | No | No | Likely | No | No |

<sup>1</sup> For language-related claims and design/analysis questions, we assessed only the potato-CRC questions.
<sup>2</sup> Touting the importance of a statistically significant difference despite the estimated effect size being scientifically insignificant.
<sup>3</sup> Touting the importance of an estimated effect size despite no statistically significant difference.
<sup>4</sup> Our assessments are based on summary data and the descriptions of the methods in all but two cases (Flood et al. and Partula et al.). We would need the raw data to definitively confirm.
<sup>5</sup> Differences in Nominal Significance (DINS) errors are when p-values from within-group tests are compared to reach statistical conclusions, which is not a valid approach.

## Conclusion

Research on the relationship between potato consumption and CRC is insufficiently reproducible and verifiable. Of the eighteen studies included in our sample, none of them publicly shared data, and we were only able to access data to reproduce studies in 2 of 18 cases (11%), and for one of those we had to leave our country to do so. These results are in line with existing observations. A meta-analysis of meta-research studies of medical and health research found that 8% (95% CI, 5%-11%) of studies declared public data availability, and 2% (95% CI, 1%-3%) actually publicly shared data (42). Gabelica et al. requested data from authors of nearly 1800 publications in which they declared data available on reasonable request, and received data in 6.8% of cases (43).

One of the publications in our set (6%) was reproducible and verifiable. This is lower than other estimates; however, other studies use varying sampling methods and definitions of reproducibility. In samples from journals *Psychological Science* and *Cognition*, with author involvement, 24% and 62% of outcomes were reproducible (6, 7). The earliest publication date for included studies was 2014, and the authors defined a major numerical error as a 10% or greater error. In another analysis of the reproducibility of primary outcomes from randomized controlled trials published in *The BMJ* and *PLOS Medicine* between 2013 and 2016, data were obtained and reproduced fully for 46% of trials (11). In a sample of papers published in *Science* in 2011-2012, an estimated 26% were reproducible (8). In 2015 publications in so-called ‘political behaviorist’ journals, 22% could be fully reproduced (12). Of a survey of registered reports in psychology literature published between 2014 and 2018, the main results were reproduced for 34%, though authors were not contacted if data or code were not publicly available (13). In social learning literature published between 1955 and 2018, 23% were reproducible (14), and among 13 economics journals published between 2008 and 2013, key results were reproducible with assistance from authors in 43% of cases (9).

One reason for the lower rate of reproducibility in our case is the low rate of data sharing. In our sample, 28% of articles were published in the 1990s. Minocher et al. showed that in social learning research, there is an decline in data recovery by half every 6 years (14). Whereas we attempted to contact authors using several methods, it is likely that some never received our communications because they are no longer active in research or have moved institutions. Nonetheless, this highlights the importance of public data sharing practices, or at minimum, long-term plans to maintain access to datasets. A second reason for low data sharing may be the observational study designs in our sample. Indeed, it would be difficult to experimentally manipulate potato consumption for the timespan needed to observe a change in CRC incidence. Our findings emphasize the lag in data sharing by observational studies behind clinical trials (44).

There are limitations to our work. We did not preregister our search or analytic plans due to the exploratory nature of our procedure development, but this is an element of rigor that should be done to increase transparency. We assessed rigor in four different ways, as each tool describes something different (risk of bias from methodological aspects of each study, a minimal set of what should be reported to interpret each study, a measure of quality, and our own additional criteria (preregistration, data and code sharing, etc.)). Each tool has strengths and limitations and inherent subjectivity, and there may be disagreement about how to interpret the results. Even more tools such as GRADE (45) could be applied. Future work could standardize the use of these tools for the goals herein. Finally, within each publication, we only attempted to reproduce analyses for potato-CRC associations, and thus cannot rule out errors in other aspects of the work.

In conclusion, for research on the relationship between potatoes and CRC, improvements can be made to increase transparency through preregistration, data and code sharing, and to increase the reporting of key methodological aspects of the study designs. Existing study designs have common weaknesses in their ability to provide causality, in the measurement of potato consumption, and in their handling of missing data. The reproducibility and verifiability of this research is not adequate and therefore reduces the trustworthiness of its findings. To our knowledge, ours is the first survey of the reproducibility and verifiability of a specific research question, and the first of the same in nutrition research. Additional work should explore this in other areas of nutrition.

## Abbreviations

CRC: Colorectal cancer
HR: Hazard Ratio
95% CI: 95% confidence interval

## Acknowledgements

We are grateful to librarian Thea Atwood at Indiana University for her assistance with search string translation. We thank Lilian Golzarri Arroyo for reviewing our decisions regarding the potential clustering issues in the papers.

## Authors’ contributions

CJV, YJ-N, BF, DBA designed research; CJV, YJ-N, PK, BO, JM, LS, KS, SR, XT, BH, SLD conducted research; CJV, YJ-N, PK, BO, BH, SLD analyzed data or performed statistical analysis; CJV, YJ-N, DBA wrote paper; DBA had primary responsibility for final content. All authors critically reviewed, edited, and approved the final manuscript.

## Data Availability

Data and code are publicly available here: https://osf.io/6jkwy/. The two datasets we received can be requested from the corresponding authors of the respective papers. We did not have any special access privileges that others would not have.

## Footnotes

1 Definition of rigor: “the strict application of the scientific method to ensure unbiased and well-controlled experimental design, methodology, analysis, interpretation and reporting of results” (1)

2 Definition of reproducibility: “obtaining consistent results using the same input data, computational steps, methods, and code, and conditions of analysis. This definition is synonymous with ‘computational reproducibility’” (1)

3 Definition of verifiability: “Verifiability subsumes, but goes beyond, reproducibility. That is, a study is said to be verifiable, and to have been verified, when: (a) the study is reproducible, and the results have been reproduced (by the definition of reproducibility above); and (b) a determination is made that the methods used to generate the results reproduced are valid methods and that the interpretations validly and logically follow from the obtained results.” (2)

