## Appendix 1-2 for "Evaluating the Reproducibility and Verifiability of Nutrition Research: A Case Study of Studies Assessing the Relationship Between Potatoes and Colorectal Cancer"

PubMed Search String

potato*[tiab] OR solanum tuberosum[tiab] OR french fry[tiab] OR french fries[tiab] OR potato chip*[tiab]

colon[tiab] OR rectum[tiab] OR rectal[tiab] OR colorectal[tiab] OR colorectum[tiab] OR bowel[tiab]

prospective OR cohort OR longitudinal OR follow-up OR case-cohort OR nested case-control

(#1 AND #2 AND #3)

Scopus Search String

TITLE-ABS ( ( potato* )  OR  ( "solanum tuberosum" )  OR  ( "french fry" )  OR  ( "french fries" )  OR  ( "potato chip*" ) )

AND

TITLE-ABS ( ( colon )  OR  ( rectum )  OR  ( rectal )  OR  ( colorectal )  OR  ( colorectum )  OR  ( bowel ) )

AND

( ( prospective )  OR  ( cohort )  OR  ( longitudinal )  OR  ( follow-up )  OR  ( case-cohort )  OR  ( "nested case-control" ) )

Web of Science Search String

TS=((potato*) OR ("solanum tuberosum") OR ("french fry") OR ("french fries") OR ("potato chip*"))

AND

TS=((colon) OR (rectum) OR (rectal) OR (colorectal) OR (colorectum) OR (bowel))

AND

TS=((prospective) OR (cohort) OR (longitudinal) OR (follow-up) OR (case-cohort) OR ("nested case-control"))

**Appendix 2**

**Reproducibility analysis of Flood et al. (2002)**

This is documentation of our attempts in reproducing tables 2 and 3 from Flood et al., doi:[10.1093/ajcn/75.5.936](https://doi.org/10.1093/ajcn/75.5.936)

### 1. Sample Size

1. Flood et al 2002 indicated in the table footnotes that 45,490 participants were used in the analysis
   - If we manually use the exclusions listed in the paper, we obtain **45,494** where discrepancies seem to be due to redacted dates that prohibited identification of 4 ineligible participants.
   - We could obtain the number **45,490** for sample size after using variable ‘exc_fl’ that was specifically made to categorize the exclusions and excluding those with >16 servings of fruit and/or veggies per day.
2. One place in the text mentioned **41,323** had ‘complete’ study data which is a discrepancy from the 45,490 indicated in the table footnotes.
   - Using same criteria, we manually obtained **41,219**.

### 2. Fruit and Vegetable Intake Quintiles

3. Table 2 calls for quintiles of fruit and vegetable intake in servings per day per 1000 kJ
   - No variables appeared to indicate servings per day in the dataset we received, but it did have frequencies of food items per week:
     - Without a variable for servings, we used frequency of consumption by dividing the weekly frequencies by seven to estimate daily servings.
     - To convert kcal to kJ servings per day, 1 kcal = 4.184 kJ.
     - Divide servings per day by kJ per day, then multiply by 1000 kJ

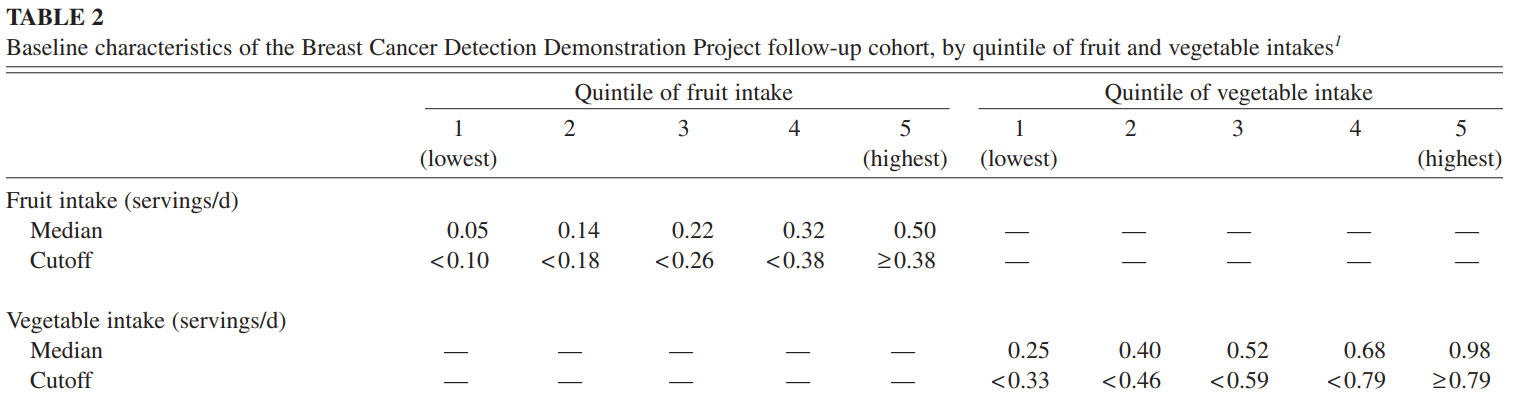

**Our Results for Median Values of Fruit and Vegetable Intake by Quintile**

| **Quintile** | **1** | **2** | **3** | **4** | **5** |
| --- | --- | --- | --- | --- | --- |
| Fruit | 0.05 | 0.14 | 0.23 | 0.33 | 0.52 |
| Vegetable | 0.21 | 0.33 | 0.43 | 0.56 | 0.81 |

**Our Results for Quintile Cutoff:**

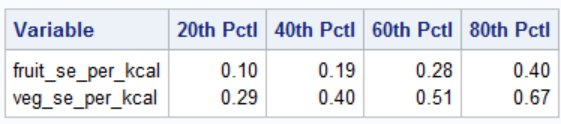

### 3. Risk models of Colorectal Cancer by F&V Quintiles

We attempted to reproduce the model Risk estimates for nutrient-density models (Table 3 “Energy-adjusted model”).

A. ***Try 1:*** Using the cutoffs defined in Table 2 of the paper:

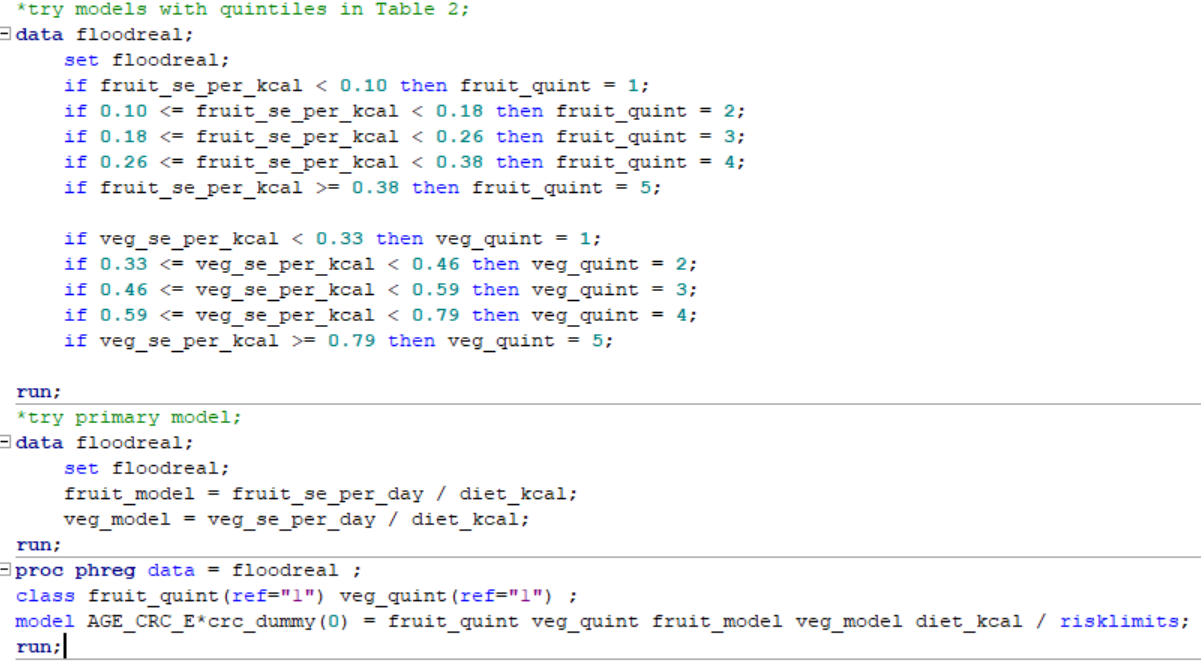

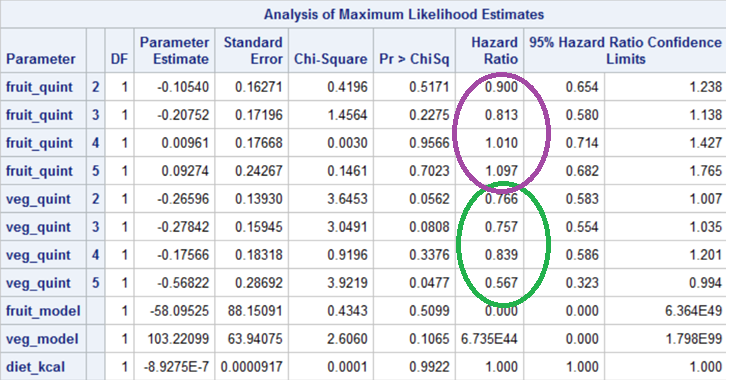

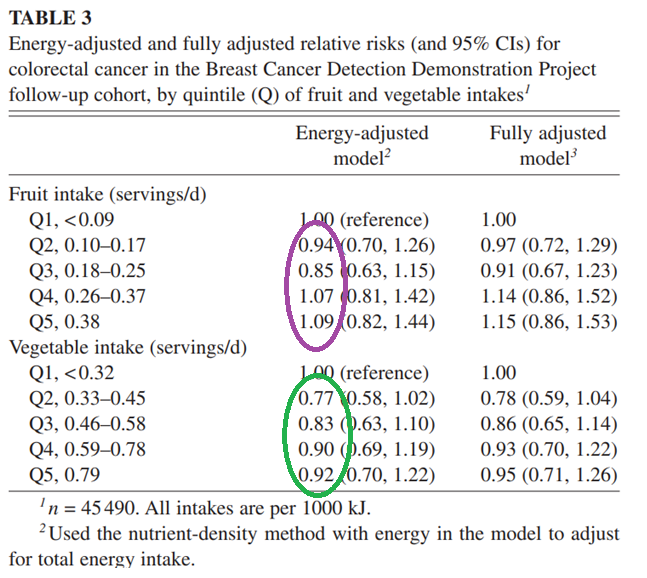

Try 1 with removing fruit_model and veg_model ^[1]^ from the model

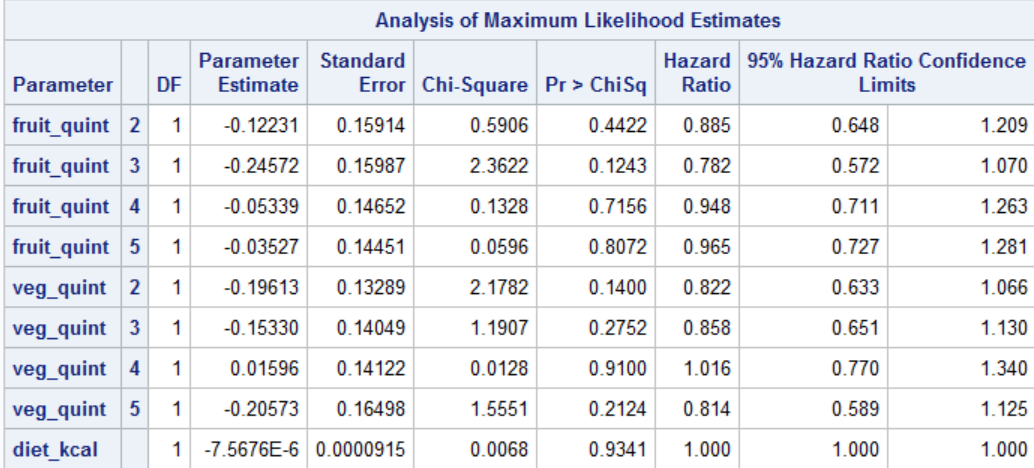

***Try 2***: Using quintile cutoffs we obtained [calculated in SAS (fruit_se_per_kcal & veg_se_per_kcal)]

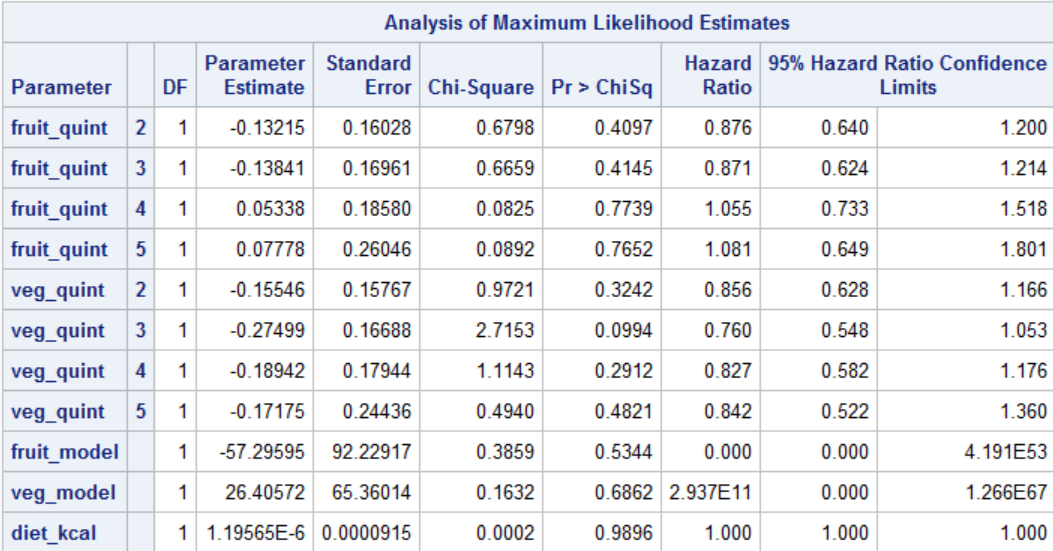

Try 2 with removing fruit_model and veg_model from model:

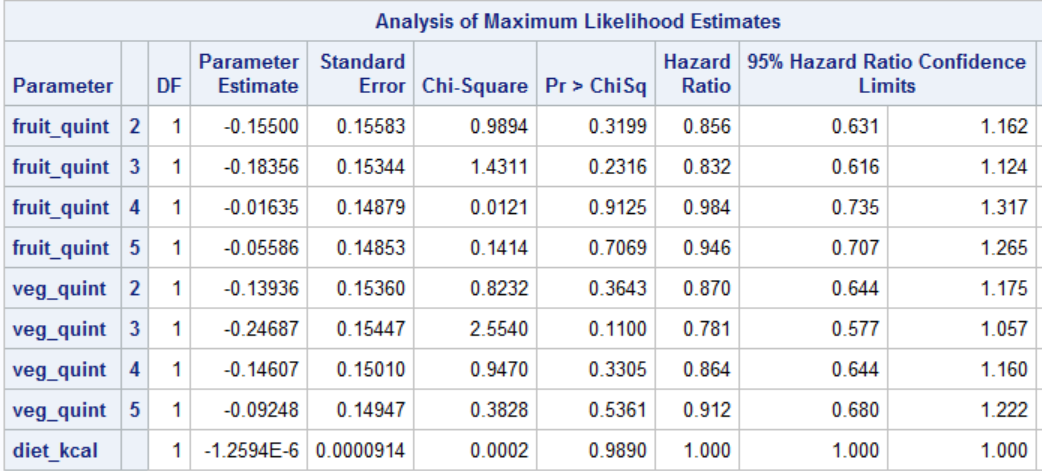

B. ***Try 3:*** Using cutoffs defined in Table 3 of paper:
   - Note that there is an internal inconsistency in the paper which appears to define the cutoffs slightly differently in Table 2 and Table 3.

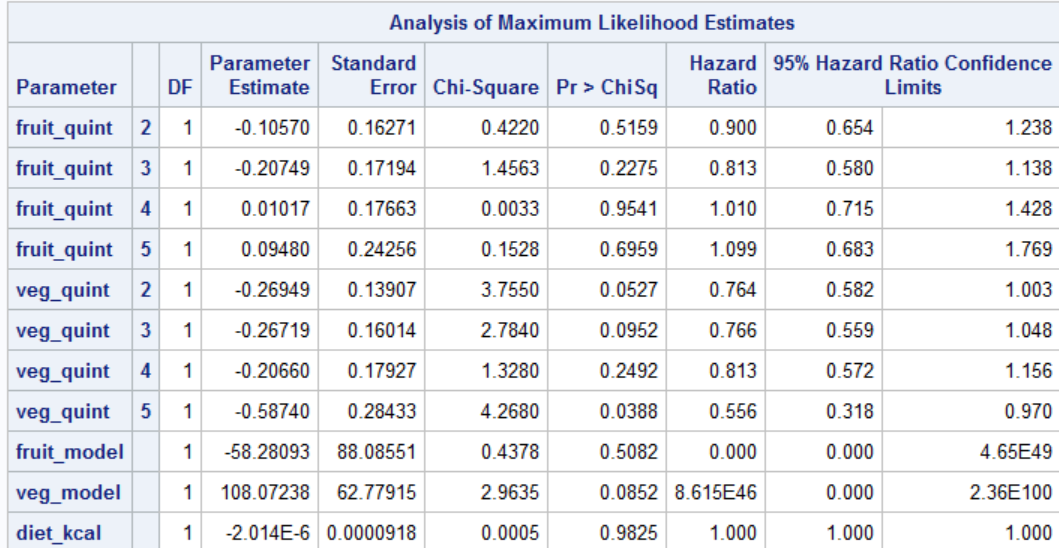

Try 3 with removing fruit_model and veg_model from the model

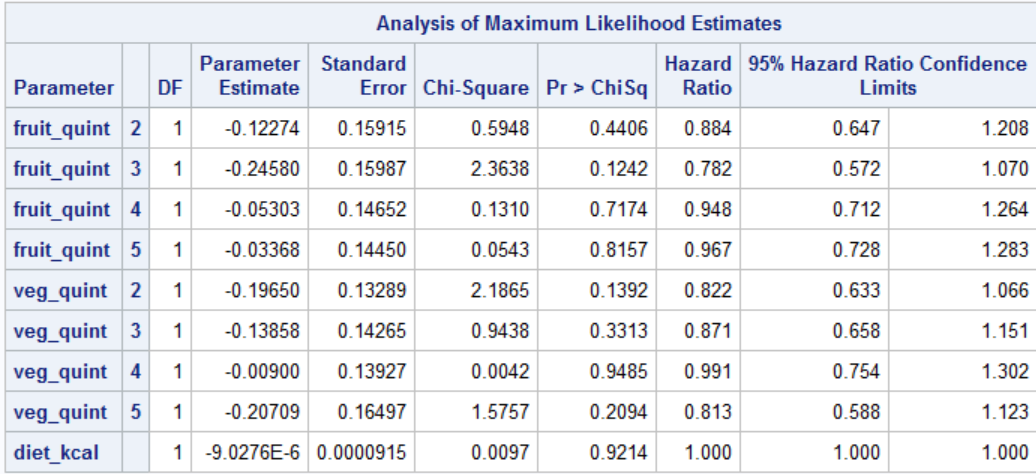

- In all three attempts, we obtained results in the same ball-park as those in the table (for fruit), but not for vegetables and for RRs in Table 3.
- Note that we obtain Hazard Ratios which are standard from the Survival analysis. We are not confident if these have been converted to Risk Ratios (RRs) in the paper, or they mean HRs.
- We also are not confident whether Flood et al. ran separate models for fruits and vegetables or included in the same model. We tried either way, and did not obtain the same numerical values as those published.

1. It is mentioned in the original paper by Flood et al. that "the primary model was the multivariate nutrient-density model, in which servings of fruit or vegetables divided by total energy entered the model along with total energy (as a separate covariate)." fruit_model is the servings of fruit divided by the total energy in kilocalories and the veg_model is the servings of veggies divided by kilocalories. As including the servings of fruits and vegetables (divided by calories) in addition to the quintiles which were also defined by servings of fruits and vegetables in the model was an unfamiliar approach to us, we did an exploratory analysis without fruit_model and veg_model variables.
